# Algorithmic Fairness of Machine Learning Models for Alzheimer’s Disease Progression Prediction

**DOI:** 10.1101/2023.07.06.23292322

**Authors:** Chenxi Yuan, Kristin A. Linn, Rebecca A. Hubbard, Alzheimer’s Disease Neuroimaging Initiative

**Affiliations:** Department of Biostatistics, Epidemiology, and Informatics, Perelman School of Medicine, University of Pennsylvania, Philadelphia, USA

**Author notes:** Authors contributed equally (Kristin A. Linn, Rebecca A. Hubbard). Correspondence: Dr. Kristin A. Linn and Dr. Rebecca A. Hubbard. Kristin A. Linn: Department of Biostatistics, Epidemiology, and Informatics, Perelman School of Medicine, University of Pennsylvania; Rebecca A. Hubbard: Department of Biostatistics, Epidemiology, and Informatics, Perelman School of Medicine, University of Pennsylvania;. Data used in preparation of this article were obtained from the Alzheimer’s Disease Neuroimaging Initiative (ADNI) database (adni.loni.usc.edu). As such, the investigators within the ADNI contributed to the design and implementation of ADNI and/or provided data but did not participate in analysis or writing of this report. A complete listing of ADNI investigators can be found at: http://adni.loni.usc.edu/wp-content/uploads/how_to_apply/ADNI_Acknowledgement_List.pdf.

**Keywords:** Alzheimer’s disease, disease progression, algorithmic fairness, machine learning, health disparities

## Abstract

**Introduction:** Alzheimer’s disease (AD) disproportionately affects older adults from marginalized communities. Predictive models using machine learning (ML) techniques have potential to improve early detection and management of AD. However, ML models potentially suffer from biases and may perpetuate or exacerbate existing disparities.

**Methods:** We investigated algorithmic fairness of logistic regression, support vector machines and recurrent neural networks for predicting progression to mild cognitive impairment and AD. Fairness was quantified across gender, ethnicity, and race subgroups using three measures: equal opportunity, equalized odds and demographic parity.

**Results:** All three ML models performed well in aggregate but demonstrated disparate performance across race and ethnicity subgroups. Compared to Non-Hispanic participants, sensitivity for predicting progression to mild cognitive impairment and to AD was 5%-9.6% and 16.8%-24.9% lower, respectively, for Hispanic participants. Sensitivity was similarly lower for Black and Asian participants compared to Non-Hispanic White participants. Models generally satisfied metrics of fairness with respect to gender.

**Discussion:** Although accurate in aggregate, models failed to satisfy fairness metrics. Fairness should be considered in the development and deployment of ML models for AD progression.

## INTRODUCTION

The development and deployment of machine learning (ML) algorithms in healthcare have received a surge of attention in recent years.^1^ ML methods are used to analyze large health datasets and have shown considerable promise across a variety of applications. Examples of successful ML applications in healthcare include disease classification using imaging data,^2, 3^ prediction of disease progression incorporating multi-modal data,^4–6^ novel biomarker discovery,^7–10^ drug repurposing^11, 12^ and characterizing disease heterogeneity.^13^ Although ML algorithms can inform clinical decision-making and potentially improve population health,^14^ there is growing concern that ML may inadvertently introduce bias into decision-making processes.^15–17^ ML algorithms may unintentionally discriminate against underrepresented and disadvantaged populations because they replicate and amplify biases in medical datasets.^1^ This impact may be the consequence of unfairness in historical and current care access and delivery, underrepresentation in clinical datasets, the use of biased or mis-specified proxy outcomes, and differences in the accessibility, usability, and effectiveness of predictive models across different groups.^18^

Since algorithms are vulnerable to biases that render their decisions unfair, *fairness*, in the context of decision-making, is the absence of any prejudice or favoritism toward an individual or group based on their inherent or acquired characteristics. An unfair algorithm skews benefits toward a particular group of people, also referred to as algorithmic bias, with respect to protected attributes.^19^ Protected attributes are features that may not be used as the basis for decisions. There is no one universal set of protected attributes. They are determined based on laws, regulations, or other policies governing a particular application domain in a particular jurisdiction. Attributes such as race, color, age, gender, national origin, religion and marital status are commonly considered *protected attributes*.^19–21^

A large body of research has been conducted on algorithmic bias in health and medicine.^22–25^ One study found algorithmic bias arising in healthcare cost prediction due to Black patients’ historically lower healthcare costs than white patients.^26^ Algorithmic underdiagnosis in chest X-ray pathology classification showed considerable disparities in automated diagnoses across ethnic and other demographic groups.^27^ A study showing that gender-biased datasets produce models that perform better for the majority class proved that ML models can spread data biases.^28^ Additionally, the Department of Health and Human Services (DHHS) has mandated identification of sources of bias and discriminatory outputs in ML algorithms^29^. However, the problem of algorithmic bias in the context of ML for Alzheimer’s disease (AD), such as the prediction of AD progression using ML approaches, has received little attention.

In this study, we characterized algorithmic fairness of longitudinal prediction models for AD progression. Using publicly available data from the Alzheimer’s Disease Neuroimaging Initiative (ADNI),^30^ we audited the fairness of three ML models of progression to AD. The overall goals of this work are to: 1) introduce and define fairness metrics relevant to models for predicting AD progression and 2) illustrate how ML algorithms can be analyzed to reveal potential disparities across protected attributes. An illustration of our model pipeline is presented in Supplementary Figure 1.

## METHODS

### Population

Data were provided by the TADPOLE challenge,^31^ derived from the Alzheimer’s Disease Neuroimaging Initiative (ADNI).^30^ ADNI was initiated in 2003 to facilitate study of AD progression. In brief, ADNI enrolled participants between the ages of 55 and 90 at 57 sites in the United States and Canada. Our dataset incorporated longitudinal data from multiple ADNI study phases and included measurements from every participant contributing data on at least two visits between September 2005 and May 2017. Clinical status at each visit was classified as: cognitively normal (CN), mild cognitive impairment (MCI) or AD. Predictor variables incorporated in our analyses included neuropsychological test scores, anatomical features from T1 magnetic resonance imaging (MRI), measures extracted from positron emission tomography (PET), and cerebrospinal fluid (CSF) biomarkers. We defined progression trajectories as transition from baseline CN to MCI, baseline MCI to AD, CN-stable and MCI-stable (patients recorded as same stage at baseline and last visit).

### Protected Attributes

To evaluate fairness criteria, subgroups were defined on the basis of demographic attributes. We focused on attributes of gender, ethnicity, and race. These attributes were chosen because previous studies in the fairness literature have highlighted algorithmic bias according to these characteristics.^28, 32^ All characteristics were classified according to participant self-report. Gender was classified as Female or Male. Ethnicity was classified as Not Hispanic/Latino or Hispanic/Latino. Participants reporting Unknown ethnicity were excluded from ethnicity-stratified analyses. Race included seven distinct groups: Asian, America-Indian/Alaskan Native, Black, Hawaiian/Other Pacific Islander, More than one, and White. We aggregated America-Indian/Alaskan Native, Hawaiian/Other Pacific Islander, More than one, and Unknown into a category labeled Other and evaluated fairness across four racial categories: Asian, Black, White, and Other.

### Study Design

We defined unfairness, or algorithmic bias, as differences in predictive performance of an ML algorithm across subpopulations defined by a protected attribute. For example, differences in sensitivity of a model for predicting AD progression in a Black population compared with a White population would be indicative of unfairness. Commonly used fairness metrics include equalized odds, equal opportunity, demographic parity, and counterfactual fairness.^19, 33^ We focused on three fairness metrics: equalized odds, equal opportunity, and demographic parity. These criteria have natural interpretations in the context of AD progression prediction. Equal opportunity is defined as equal sensitivity or true positive rates (TPR) of the ML algorithm across all levels of the protected attribute.^34^ An AD progression algorithm would exhibit equal opportunity if individuals who truly did progress to AD were equally likely to be identified by the algorithm across all protected groups. Equalized odds requires that an algorithm exhibit equal opportunity and equal specificity or false positive rates (FPR) across groups. Demographic parity, also known as statistical parity, is the equivalence of a predicted event’s probability across sensitive attribute groups.^33^ In our AD case study, demographic parity with respect to gender would be satisfied if females and males were predicted to develop AD with equal probability. When real differences in outcome prevalence exist across groups, achieving demographic parity may be undesirable. Importantly, unless prevalence is equal across subgroups it is impossible to simultaneously satisfy all metrics. Table 1 presents mathematical definitions of these fairness metrics.

**Table 1.** Mathematical definitions of three common fairness metrics. *Y* is the observed outcome, *Ŷ* is a prediction of *Y*, and *A* is a binary protected attribute.

| **Fairness Metrics** | **Definition** | **In Alzheimer's Disease** |
| --- | --- | --- |
| Equal Opportunity | The True Positive Rates are the same across groups:<br>$\mathbb{P}(\hat{Y} = 1 \vert A = 0, Y = 1) = \mathbb{P}(\hat{Y} = 1 \vert A = 1, Y = 1)$ | The probability of correctly predicting that an individual progresses to AD is the same for subgroups defined by a protected attribute such as race |
| Equal Odds | The True Positive Rates and False Positive Rates are the same across groups:<br>$\mathbb{P}(\hat{Y} = 1 \vert A = 0, Y = y) = \mathbb{P}(\hat{Y} = 1 \vert A = 1, Y = y), y \in \{0, 1\}$ | The probability of correctly predicting that an individual progresses to AD and the probability of incorrectly predicting progression to AD for those who do not are the same for subgroups defined by a protected attribute such as race |
| Demographic Parity | Equal probability of being classified with the positive label<br>$\mathbb{P}(\hat{Y} = 1 \vert A = 0) = \mathbb{P}(\hat{Y} = 1 \vert A = 1)$ | The proportion of individuals predicted to progress to AD is the same across subgroups defined by a protected attribute such as race |

### Prediction Models

We assessed fairness with respect to the task of predicting AD progression with ML algorithms.^35^ We selected three ML models for evaluation in this study: logistic regression (LR), support-vector machine (SVM), and recurrent neural networks (RNN). We included LR and SVM because they are well-established ML models commonly used for prediction problems and are often presented as comparators for new models.^36–38^ As a deep learning model, RNN has shown promise in on the AD progression domain^39, 40^ and has been applied to prediction problems,^37, 41^ demonstrating great improvement over other ML models on prediction accuracy. The RNN model we tested in this study is from Nguyen.^35^ Models were trained using the ADNI dataset to predict participants’ clinical status at the subsequent visit as CN, MCI, or AD. More specifically, given the data collected for a subject at baseline, the models predicted the diagnosis stage at subsequent time points. Additional details of model implementation and training are provided in the Online Supplement.

We used cross-validation for model selection and evaluation. The data were randomly partitioned into 10 equal subsets. In each iteration of the 10-fold cross-validation, 80% were used for training, 10% were used for model validation, and the remaining 10% were included in the test set. In each iteration, the training set was used for model fitting, the validation set was used to select values for hyperparameters, and the test set was used to evaluate the model’s performance under the optimal set of hyperparameters identified by the validation set. All continuous variables were *z*-normalized using the training set to estimate the mean and standard deviation, which were then utilized to z-normalize the validation and test sets.

### Fairness Analysis

To assess algorithmic fairness, we calculated fairness metrics on each of the 10 test sets using predictions from each model. All metrics are reported as the mean and standard deviation across the 10 values. Evaluations were conducted separately by demographic group. We first assessed equal opportunity by computing the TPR for sub-groups defined by each protected attribute separately for each cognitive functioning trajectory (i.e. CN to MCI, MCI to AD, stable CN, stable MCI) and each of the three models. The TPR quantifies the proportion of individuals experiencing a given trajectory who are correctly predicted to follow that trajectory. For instance, TPR of CN to MCI represents the probability of correctly predicting that an individual progresses from CN to MCI. A TPR value of 1 indicates that the model has achieved perfect sensitivity in identifying the positive instances within the particular category. If the TPRs for each trajectory are similar across protected feature categories, such a result suggests the model attains equal opportunity. We also calculated differences in TPR between subgroups for each protected attribute. In addition to TPR, we calculated the FPR. Specifically, for a given trajectory the FPR is defined as the proportion of individuals who did not experience that trajectory who are incorrectly predicted to follow that trajectory. For example, FPR of CN to MCI represents the probability of predicting progression from CN to MCI for an individual who, in reality, does not progress. An algorithm must demonstrate both equal TPR and equal FPR across subgroups to satisfy the equalized odds criterion. To assess demographic parity, we computed the predicted probability for each trajectory and each demographic subgroup. We report the difference in this predicted probability across subgroups for each sensitive attribute, trajectory, and ML model. Finally, we also calculated the empirical probability of each trajectory stratified by demographic subgroup. Hypothesis testing was not employed in this context due to the lack of independence among predicted values on the test set^42^. Additionally, due to small sample sizes among some subgroups, hypothesis testing is expected to have low power. We therefore focus on point estimation and interpretation of point estimates in light of their variability. An overview of the experimental procedure was shown in Figure 1.

**Figure 1.**
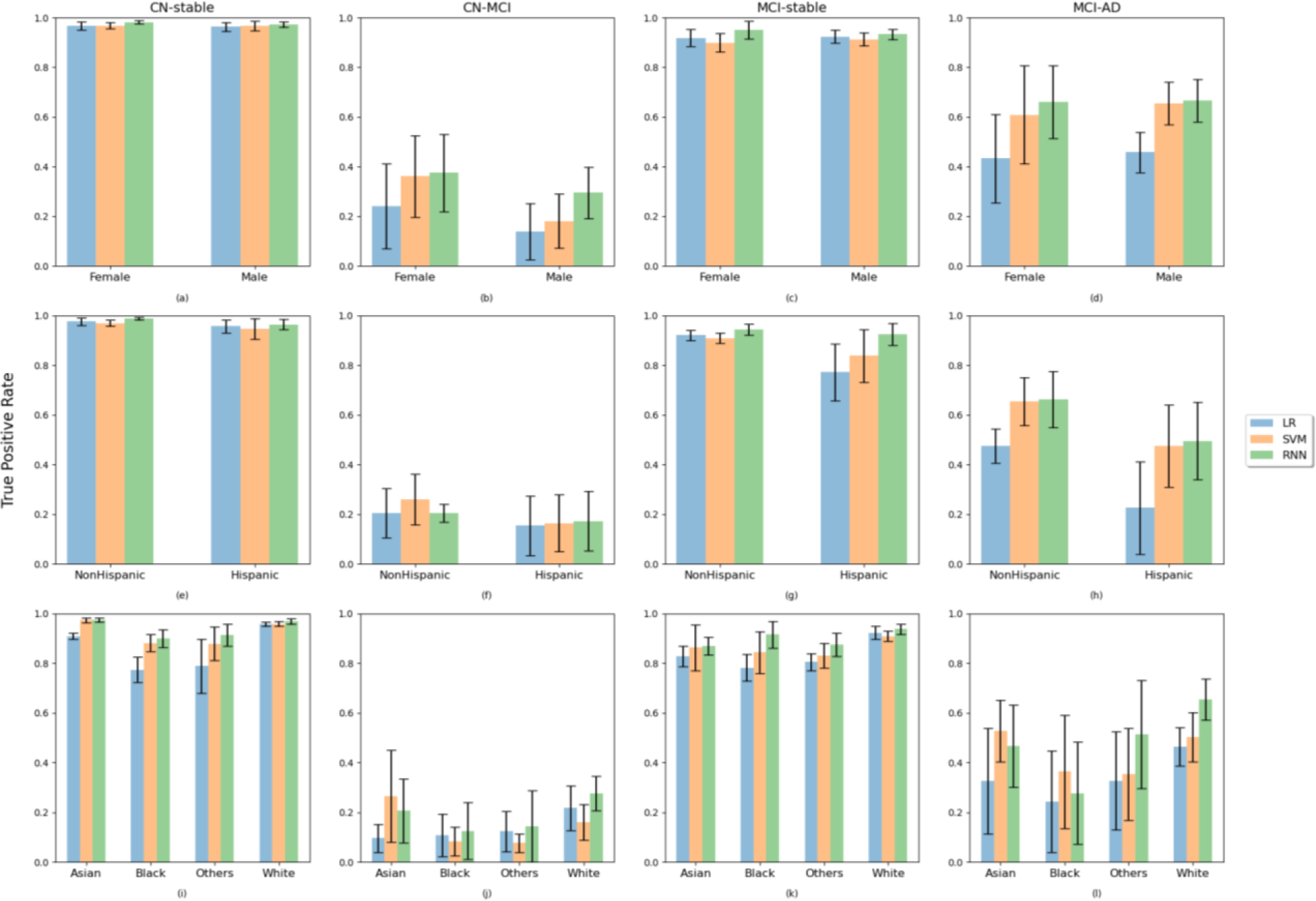
Comparison of True Positive Rates across subgroups of gender, ethnicity and race for three models. The results are averaged over 10 test sets using predictions from the LR, SVM, and RNN models. Bars present the mean values across 10 test sets and error bars represent the standard deviation of the 10 mean values.

## RESULTS

### Study cohort

The dataset included 1730 subjects aged 54 to 91 years, each scanned at multiple timepoints, contributing an average of 7.3 (standard deviation [SD] 4.0) observations per participant over an average of 3.6 (SD 2.5) years. The distribution of participant characteristics stratified by clinical status at the baseline and last visit is provided in Table 2. Backward transitions (i.e., MCI to CN, AD to MCI or CN) and transition from CN to AD were rarely observed (Supplementary Table S1) and were, therefore, not included in fairness evaluations. Aggregate performance of the ML models was good (Supplementary Table S2) and similar to published results ^35^.

**Table 2.**
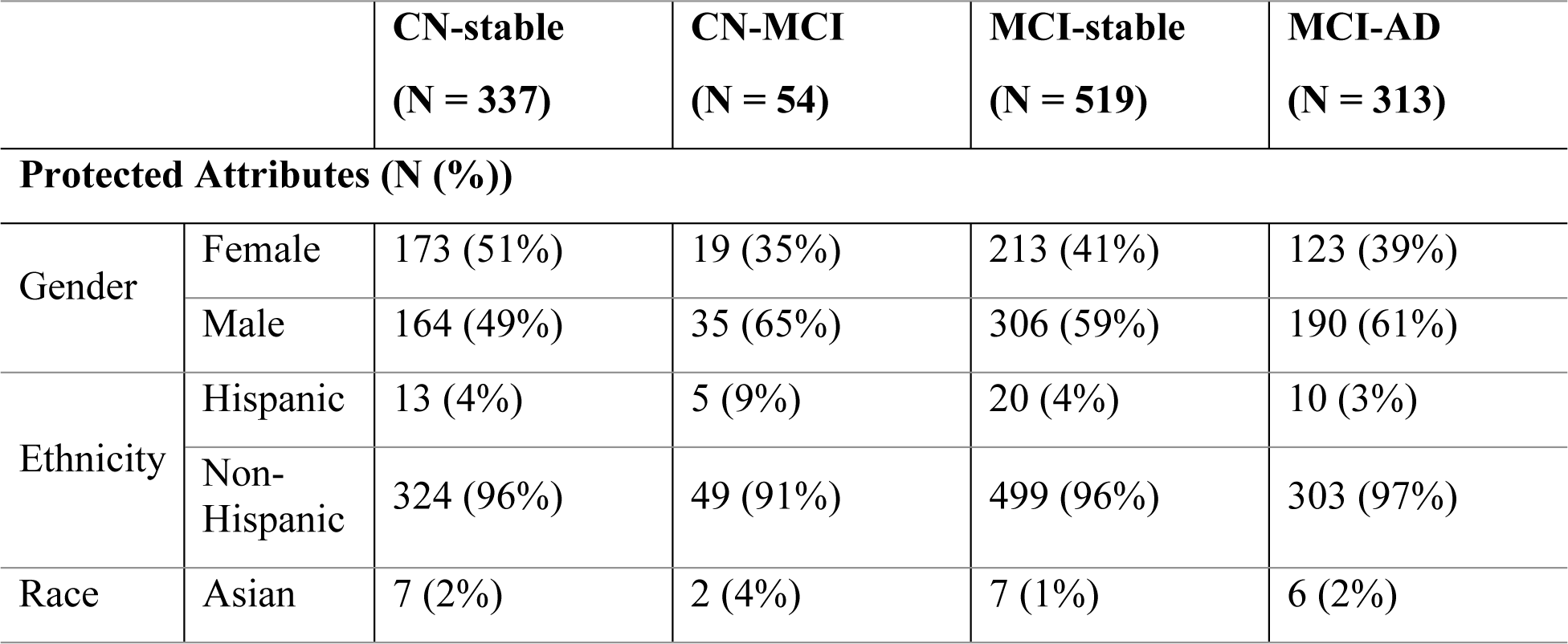
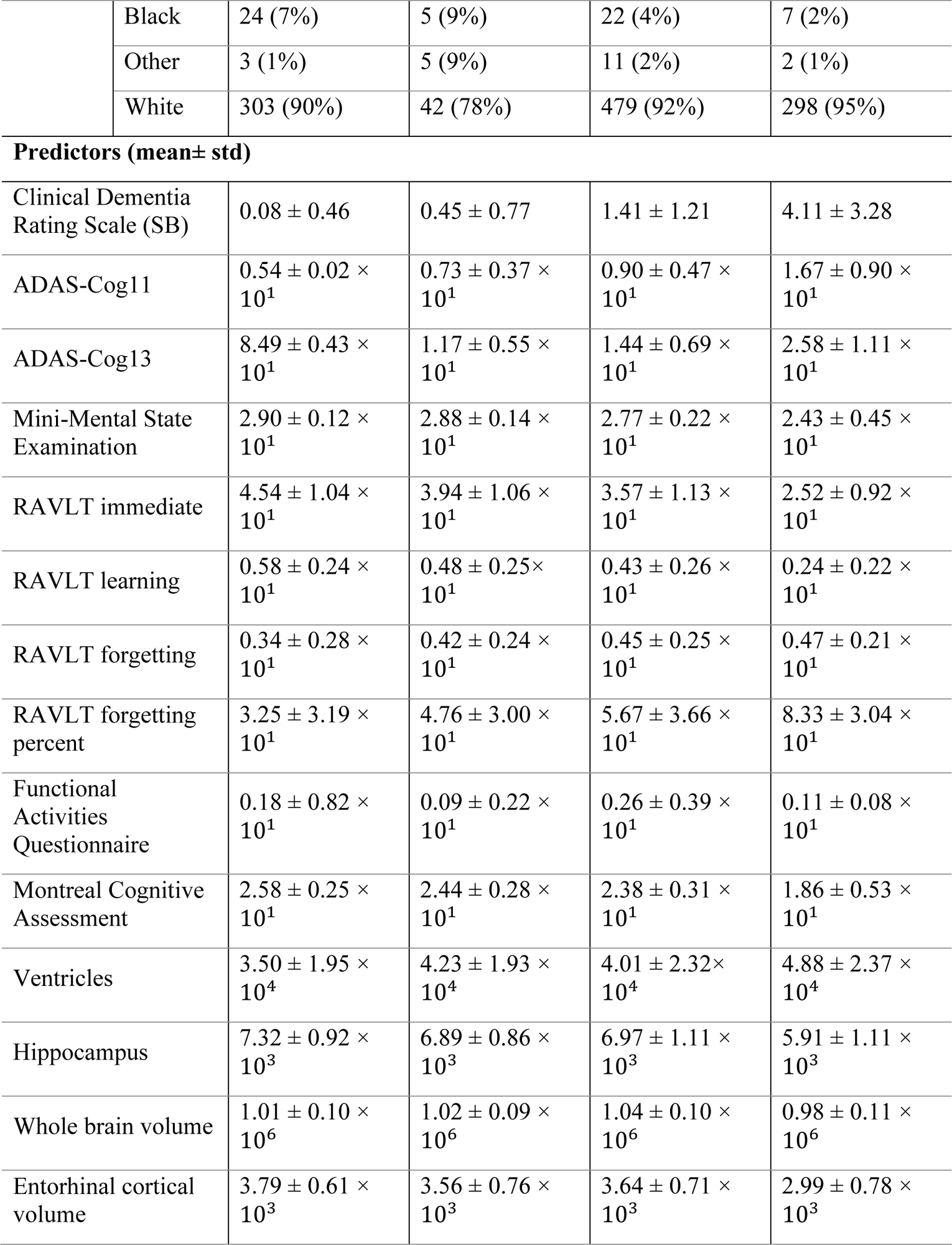
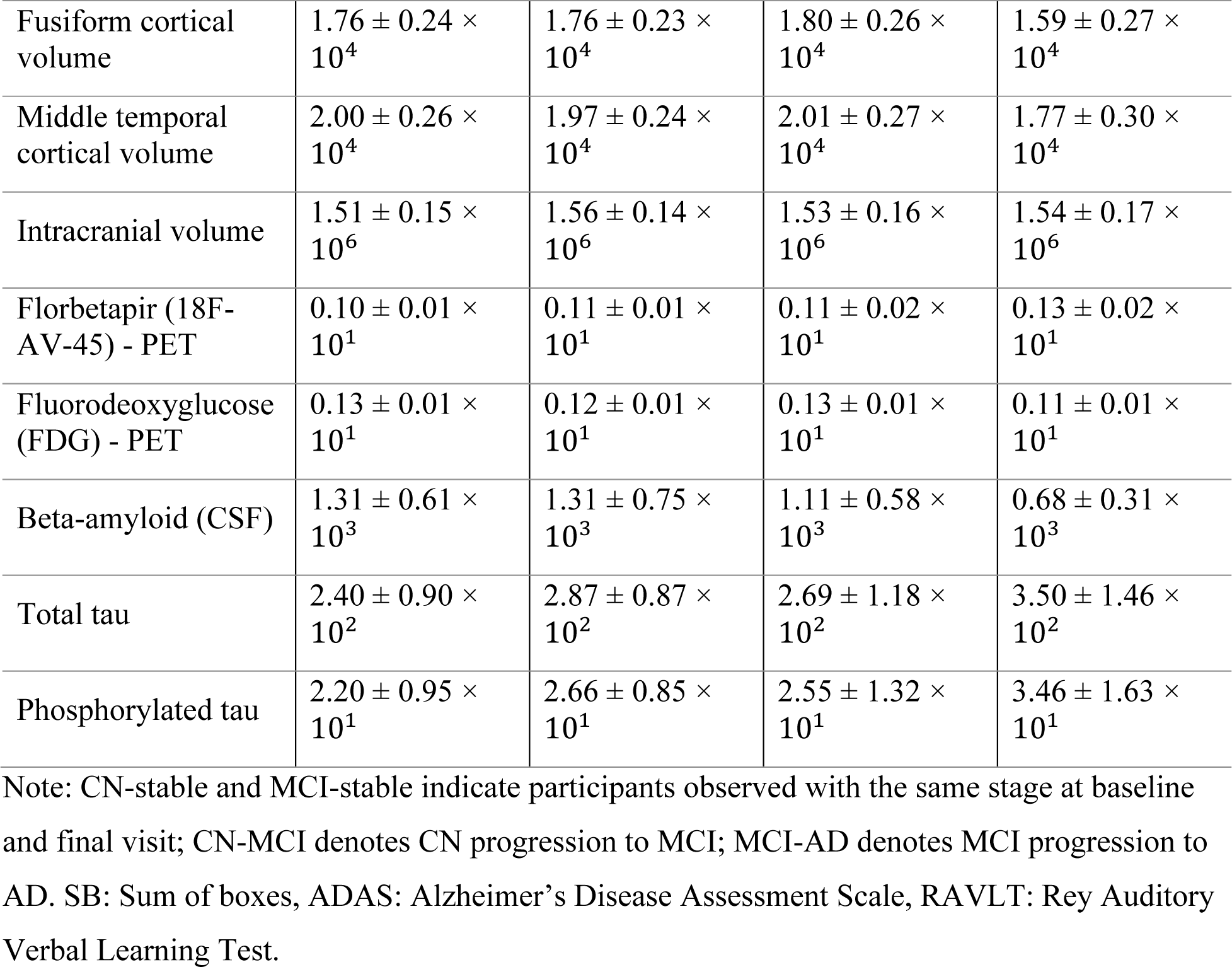
Summary statistics for protected attributes and predictor variables stratified by cognitive functioning trajectory.

|  |  | **CN-stable**<br>(N = 337) | **CN-MCI**<br>(N = 54) | **MCI-stable**<br>(N = 519) | **MCI-AD**<br>(N = 313) |
| --- | --- | --- | --- | --- | --- |
| **Protected Attributes (N (%))** |  |  |  |  |  |
| Gender | Female | 173 (51%) | 19 (35%) | 213 (41%) | 123 (39%) |
|  | Male | 164 (49%) | 35 (65%) | 306 (59%) | 190 (61%) |
| Ethnicity | Hispanic | 13 (4%) | 5 (9%) | 20 (4%) | 10 (3%) |
|  | Non-Hispanic | 324 (96%) | 49 (91%) | 499 (96%) | 303 (97%) |
| Race | Asian | 7 (2%) | 2 (4%) | 7 (1%) | 6 (2%) |
|  | Black | 24 (7%) | 5 (9%) | 22 (4%) | 7 (2%) |
|  | Other | 3 (1%) | 5 (9%) | 11 (2%) | 2 (1%) |
|  | White | 303 (90%) | 42 (78%) | 479 (92%) | 298 (95%) |

**Predictors (mean ± std)**
|  |  |  |  |  |
| --- | --- | --- | --- | --- |
| Clinical Dementia Rating Scale (SB) | 0.08 ± 0.46 | 0.45 ± 0.77 | 1.41 ± 1.21 | 4.11 ± 3.28 |
| ADAS-Cog11 | 0.54 ± 0.02 $\times 10^1$ | 0.73 ± 0.37 $\times 10^1$ | 0.90 ± 0.47 $\times 10^1$ | 1.67 ± 0.90 $\times 10^1$ |
| ADAS-Cog13 | 8.49 ± 0.43 $\times 10^1$ | 1.17 ± 0.55 $\times 10^1$ | 1.44 ± 0.69 $\times 10^1$ | 2.58 ± 1.11 $\times 10^1$ |
| Mini-Mental State Examination | 2.90 ± 0.12 $\times 10^1$ | 2.88 ± 0.14 $\times 10^1$ | 2.77 ± 0.22 $\times 10^1$ | 2.43 ± 0.45 $\times 10^1$ |
| RAVLT immediate | 4.54 ± 1.04 $\times 10^1$ | 3.94 ± 1.06 $\times 10^1$ | 3.57 ± 1.13 $\times 10^1$ | 2.52 ± 0.92 $\times 10^1$ |
| RAVLT learning | 0.58 ± 0.24 $\times 10^1$ | 0.48 ± 0.25 $\times 10^1$ | 0.43 ± 0.26 $\times 10^1$ | 0.24 ± 0.22 $\times 10^1$ |
| RAVLT forgetting | 0.34 ± 0.28 $\times 10^1$ | 0.42 ± 0.24 $\times 10^1$ | 0.45 ± 0.25 $\times 10^1$ | 0.47 ± 0.21 $\times 10^1$ |
| RAVLT forgetting percent | 3.25 ± 3.19 $\times 10^1$ | 4.76 ± 3.00 $\times 10^1$ | 5.67 ± 3.66 $\times 10^1$ | 8.33 ± 3.04 $\times 10^1$ |
| Functional Activities Questionnaire | 0.18 ± 0.82 $\times 10^1$ | 0.09 ± 0.22 $\times 10^1$ | 0.26 ± 0.39 $\times 10^1$ | 0.11 ± 0.08 $\times 10^1$ |
| Montreal Cognitive Assessment | 2.58 ± 0.25 $\times 10^1$ | 2.44 ± 0.28 $\times 10^1$ | 2.38 ± 0.31 $\times 10^1$ | 1.86 ± 0.53 $\times 10^1$ |
| Ventricles | 3.50 ± 1.95 $\times 10^4$ | 4.23 ± 1.93 $\times 10^4$ | 4.01 ± 2.32 $\times 10^4$ | 4.88 ± 2.37 $\times 10^4$ |
| Hippocampus | 7.32 ± 0.92 $\times 10^3$ | 6.89 ± 0.86 $\times 10^3$ | 6.97 ± 1.11 $\times 10^3$ | 5.91 ± 1.11 $\times 10^3$ |
| Whole brain volume | 1.01 ± 0.10 $\times 10^6$ | 1.02 ± 0.09 $\times 10^6$ | 1.04 ± 0.10 $\times 10^6$ | 0.98 ± 0.11 $\times 10^6$ |
| Entorhinal cortical volume | 3.79 ± 0.61 $\times 10^3$ | 3.56 ± 0.76 $\times 10^3$ | 3.64 ± 0.71 $\times 10^3$ | 2.99 ± 0.78 $\times 10^3$ |
| Fusiform cortical volume | $1.76 \pm 0.24 \times 10^4$ | $1.76 \pm 0.23 \times 10^4$ | $1.80 \pm 0.26 \times 10^4$ | $1.59 \pm 0.27 \times 10^4$ |
| Middle temporal cortical volume | $2.00 \pm 0.26 \times 10^4$ | $1.97 \pm 0.24 \times 10^4$ | $2.01 \pm 0.27 \times 10^4$ | $1.77 \pm 0.30 \times 10^4$ |
| Intracranial volume | $1.51 \pm 0.15 \times 10^6$ | $1.56 \pm 0.14 \times 10^6$ | $1.53 \pm 0.16 \times 10^6$ | $1.54 \pm 0.17 \times 10^6$ |
| Florbetapir (18F-AV-45) - PET | $0.10 \pm 0.01 \times 10^1$ | $0.11 \pm 0.01 \times 10^1$ | $0.11 \pm 0.02 \times 10^1$ | $0.13 \pm 0.02 \times 10^1$ |
| Fluorodeoxyglucose (FDG) - PET | $0.13 \pm 0.01 \times 10^1$ | $0.12 \pm 0.01 \times 10^1$ | $0.13 \pm 0.01 \times 10^1$ | $0.11 \pm 0.01 \times 10^1$ |
| Beta-amyloid (CSF) | $1.31 \pm 0.61 \times 10^3$ | $1.31 \pm 0.75 \times 10^3$ | $1.11 \pm 0.58 \times 10^3$ | $0.68 \pm 0.31 \times 10^3$ |
| Total tau | $2.40 \pm 0.90 \times 10^2$ | $2.87 \pm 0.87 \times 10^2$ | $2.69 \pm 1.18 \times 10^2$ | $3.50 \pm 1.46 \times 10^2$ |
| Phosphorylated tau | $2.20 \pm 0.95 \times 10^1$ | $2.66 \pm 0.85 \times 10^1$ | $2.55 \pm 1.32 \times 10^1$ | $3.46 \pm 1.63 \times 10^1$ |

Note: CN-stable and MCI-stable indicate participants observed with the same stage at baseline and final visit; CN-MCI denotes CN progression to MCI; MCI-AD denotes MCI progression to AD. SB: Sum of boxes, ADAS: Alzheimer’s Disease Assessment Scale, RAVLT: Rey Auditory Verbal Learning Test.

### Equal Opportunity and Equalized Odds

Figure 1(a-d) shows TPR for the four progression cases stratified by gender for each of the three models. For CN-stable and MCI-stable, the TPR was close to 1, and there were no major differences in TPR between females and males. The differences in TPR between males and females were approximately 0.5% for CN-stable and ranged from 0.4% to 1.7% or MCI-stable across the three models (Supplementary Figure S2(a)). For transition from CN to MCI, there was a notable difference in TPR between genders, with all three models performing better for females than males with 10.3%, 15.0% and 10.4% absolute increases for LR, SVM and RNN, respectively. For transition from MCI to AD, small differences in TPR were observed between genders. The three models performed similarly overall, but RNN had higher TPR for predicting progression from CN to MCI and MCI to AD as well as less variability across test sets (Supplementary Figure S2(a)).

Figure 1(e-h) shows TPR for trajectory classes stratified by ethnicity. Overall, across the trajectories and models, TPR was higher for Non-Hispanic participants compared to Hispanic participants. Differences in TPR between Non-Hispanic and Hispanic participants were around 2% for CN-stable and ranged from 3% to 28% for MCI-stable across the three models (Supplementary Figure S2(b)). Differences in TPR were larger for progression from CN to MCI and MCI to AD. Specifically, TPR for Hispanic participants was approximately 5%, 9.6% and 3.2% lower than for Non-Hispanic participants for progression from CN to MCI and 24.9%, 18% and 16.8% for MCI to AD for LR, SVM and RNN respectively. In most cases, RNN had higher TPR than the other two models. Across the models for MCI to AD, RNN had the highest TPR and smallest difference in TPR between Hispanic and Non-Hispanic participants (Supplementary Figure S2(b)).

Comparisons of TPR across racial groups are shown in Figure 1(i-l). For CN-stable, TPR was high for White participants (TPR = 95.7-97.0%) and lower for other groups (TPR = 70.9-80.4%, 77.3-91.6% and 76.9-88.4%). Patterns across racial groups for MCI-stable were similar to those for CN-stable. For CN to MCI, Asian participants had higher TPR than other groups for SVM. TPR for Black participants was the lowest for CN to MCI progression (8.1-18.2% lower than Asian, 0.7-1.9% lower than Other and 7.7-15.2% lower than White). White participants had higher TPR for progression from MCI to AD for two of the three models (Supplementary Figure S2(b).

For all three models, FPR was lower for females compared to males for all trajectories (Figure 2(a-d)). Similarly, Non-Hispanic participants had lower FPR than Hispanic participants for all trajectories. For racial groups, FPR for Black participants was higher compared to other race sub-groups for CN-stable. For MCI-stable, FPR was higher for Asian participants compared to other groups. Overall, the FPR for progression from CN to MCI and MCI to AD was far lower than for stable CN and MCI. However, the large error bars for the Asian, Black and Other subgroups reflect uncertainty in the FPR point estimates due to the small sample sizes of these groups, especially in the two forward transition cases (CN to MCI and MCI to AD), making it difficult to draw conclusions. As a result, assessment of equal odds is limited for these sub-groups.

**Figure 2.**
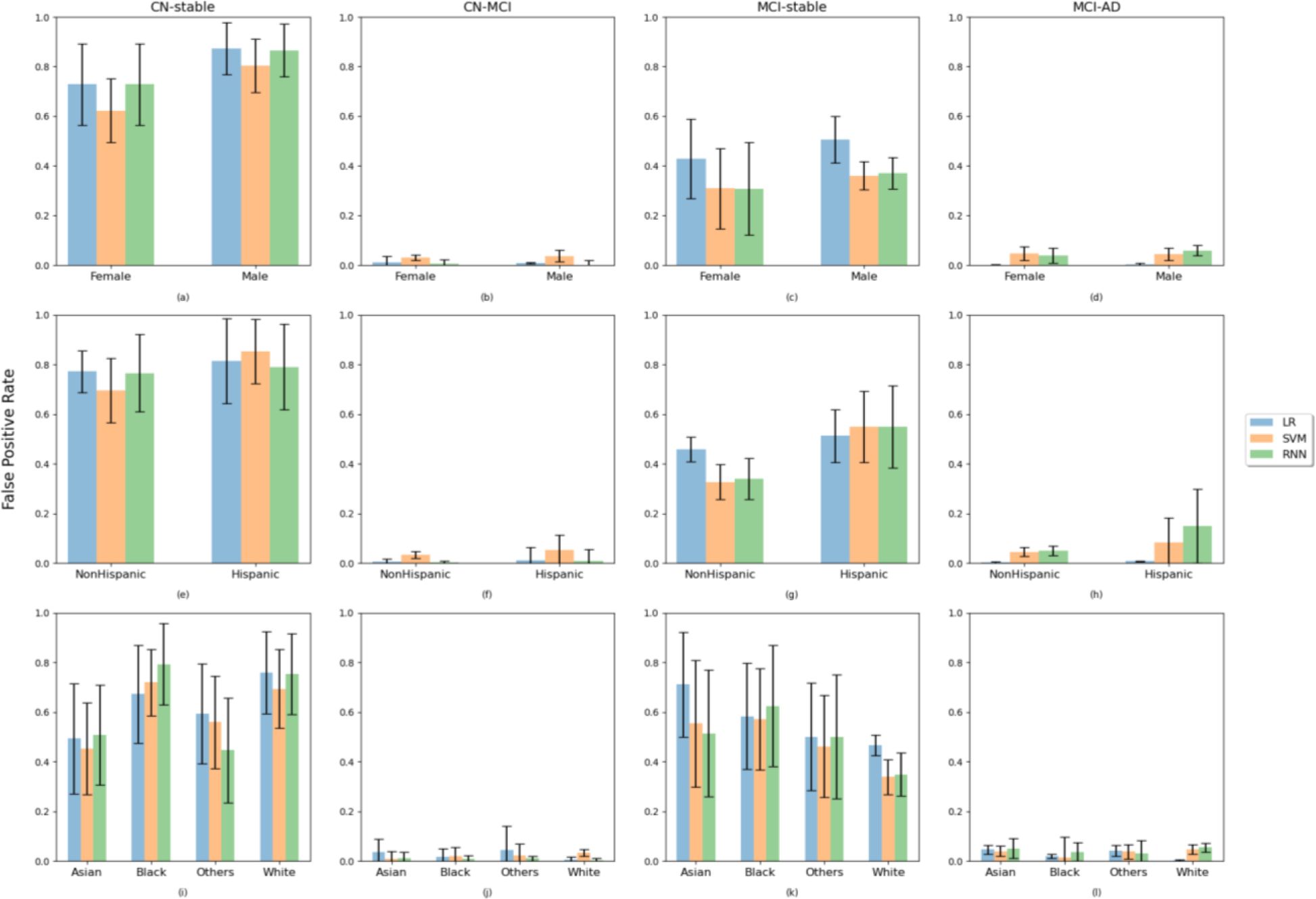
Comparison of False Positive Rates across subgroups of gender, ethnicity and race for three models. The results are averaged over 10 test sets using predictions from the LR, SVM, and RNN models. Bars present the mean values across 10 test sets and error bars represent the standard deviation of the 10 mean values.

### Demographic parity

Observed and predicted prevalence of cognitive functioning trajectories differed across groups defined by the protected attributes (Figure 3). Across all three models, the probability of being predicted to be CN-stable or MCI-stable was higher than the observed prevalence, whereas the probability of being predicted to transition from CN to MCI or MCI to AD was generally lower than or similar to the observed prevalence.

**Figure 3.**
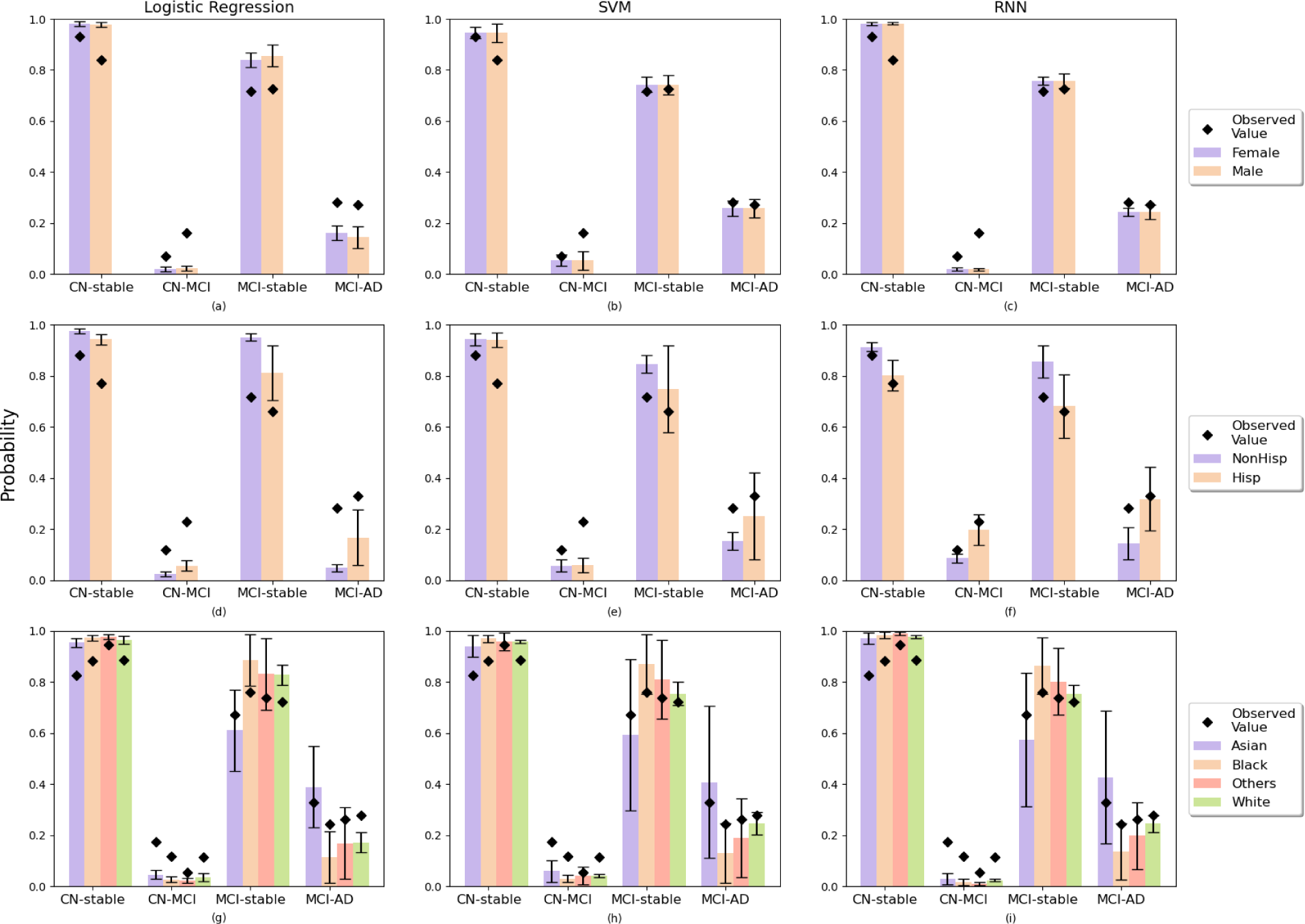
Comparison of predicted probability of progression cases across subgroups of gender, ethnicity, and race for three models. The results are averaged over 10 test sets using predictions from the LR, SVM, and RNN models. Bars represent the mean values across 10 test sets and error bars represent a corresponding standard deviation of the 10 mean values. Dots represent the average value of the empirical probability of each trajectory stratified by demographic subgroup on 10 test sets.

Female participants who were CN at baseline had slightly higher predicted probability of CN-stable. (Figure 3(a-c)), with differences ranging from 0.2% to 0.7% across three models. Conversely, the predicted probabilities of MCI-stable and MCI to AD were slightly lower for female compared to male participants, with differences ranging from 0.4% to 1.7% across the three models. These were similar to the empirical differences in prevalence between male and female participants. Notable differences between predicted and empirical probabilities were found for male participants who were CN at baseline. Specifically, the difference between predicted and observed probabilities of progressing to MCI were 13.8%, 9.8% and 13.9% for male participants, while for female participants differences were 4.8%, 1.5% and 5.0% for LR, SVM and RNN, respectively. Across all models, predictions based on the RNN were more similar to the empirical probabilities of MCI progression compared to predictions from LR and SVM (Supplementary Figure S3(a)).

The predicted probabilities of CN-stable and MCI-stable were higher for Non-Hispanic participants compared to Hispanic, consistent with the empirical distribution. Conversely, the predicted probability of progression from CN to MCI and MCI to AD for Non-Hispanic participants was lower than for Hispanic participants. The differences were 3.1%, 0.2% and 13.1% for CN progression, and 2.1%, 9.8% and 17.4% for MCI progression across LR, SVM and RNN, respectively. Discrepancies between predicted and observed probabilities for Hispanic participants were 17.2% and 31.3% for LR, 16.9% and 8.8% for SVM, and 3.1% and 2.1% for RNN, for CN progression and MCI progression respectively.

Across racial sub-groups, Asian participants had the lowest predicted probability of both CN-stable and MCI-stable, and had the highest predicted probability of progression from CN to MCI and MCI to AD. Black participants had the highest predicted probability of MCI-stable and the lowest predicted probability of progression from CN to MCI and MCI to AD. Additionally, for CN-stable and CN to MCI, the largest differences between predicted and observed values were for Asian participants with 12.8%, 11.5% and 14.4% differences for LR, SVM and RNN, respectively. For MCI-stable and progression from MCI to AD, Black participants were observed to have the largest differences between predicted and observed values (12.6% for LR, 11.2% higher for SVM, 10.5% higher for RNN). This indicates that Black participants with MCI at baseline were more likely to be misclassified as progressing to AD. In contrast, Asian participants who were CN at baseline were most likely to be misclassified as non-progressors.

## DISCUSSION

We evaluated the fairness of ML models for predicting progression of Alzheimer’s disease across sub-groups defined by gender, race, and ethnicity. Although the three models we evaluated performed well in aggregate, they failed to satisfy metrics of fairness with respect to the protected attributes we investigated. The investigation of equal opportunity, equalized odds, and demographic parity found that models exhibited little unfairness with respect to gender but had notable deficits in fairness across race and ethnicity sub-groups.

Due to differences in prevalence of progression for male and female participants, the ML models investigated did not satisfy the criterion of demographic parity with respect to gender. All three models under-predicted the probability of progressing from CN to MCI for both male and female participants, but the discrepancy between observed and predicted probabilities of progression were larger for male participants. This finding could be attributable to greater heterogeneity of trajectories in men compared to women. Progression from MCI to AD was also under-predicted by all three models. However, this under-prediction was less severe for RNN compared to the other two models.

Models displayed unfairness with respect to multiple metrics across ethnicity groups. However, uncertainty in estimates of TPR was high for Hispanic participants due to small sample sizes, making it difficult to draw firm conclusions regarding model performance for this sub-group. Notable discrepancies between observed and predicted probabilities of transition from CN to MCI were observed for Hispanic participants. These results highlight how under-representation can introduce unfairness. In the ADNI dataset only 3% of participants were Hispanic, and, consequently, models tended to perform poorly for this group. However, the deep learning model (RNN) demonstrated improved performance relative to the other two models through smaller differences between predicted and observed probabilities of progression for Hispanic participants.

Estimates of model performance for participants in the Asian, Black, and Other race groups had wide error bars due to limited sample size. Black participants in the MCI group at baseline tended to be incorrectly predicted to transition to AD. Asian participants who were CN tended to be incorrectly under-predicted to transition to MCI. A comparison of the three ML models demonstrated some improvement of the deep learning model (RNN) compared to the other models. Notably, for individuals progressing to AD and participants of Black race, RNN outperformed the other two models in the sense that the discrepancies between predicted probability and observed prevalence of AD were smaller than the other two models.

Sources of unfairness in ML models include sampling bias and implicit cultural biases that are reflected in the data. The health domain may also feature systemic biases inherent in biological processes that may not be possible to mitigate.^43^ In the AD domain, there are neuropsychiatric differences across racial and ethnic groups, some of which exist due to systemic racism, that affect disease prevalence.^26, 44, 45^ Therefore, demographic parity may not be desirable when real differences in AD disease prevalence exist. One feasible approach to evaluating fairness in this setting may be to create an adjusted demographic parity measure that incorporates a tolerance for verified differences in prevalence across protected groups.^18^

The equal opportunity and equalized odds metrics (based on TPR and FPR) are desirable criteria to satisfy because they represent equal performance accuracy of ML models across protected subgroups. However, these two metrics are limited. Equal opportunity only considers TPR and fails to encapsulate other measures of diagnostic error or value such as the positive predictive value of a model. The appropriate metric to optimize in a given context depends on the intended use case.^46^ Metrics considered in this study can help surface important normative questions about decision-making, as well as trade-offs and tensions between different potential interpretations of fairness.

Our study has several limitations. First, our study is limited to three ML models—logistic regression, support vector machines, and recurrent neural networks—trained to perform the specific task of predicting a future disease state given historical information and disease state of individuals. It is not possible to extrapolate these results to fairness for other models or prediction tasks. Second, the study reveals the existence of unfairness in AD progression prediction, but it does not identify the source of unfairness in this context or how to mitigate it. Unfairness may arise due to features of the data or algorithms, and our investigation does not distinguish between these two sources. Potential data biases include insufficient sample size in some sub-groups as well as differential misclassification of disease stage and informative missingness.^34^

Algorithmic bias arises when the bias is not present in the input data but is added purely by the algorithm.^47^ It is generated by choices in the algorithmic design including choice of optimization function, regularization, and loss function. Choices for each of these aspects of the algorithm can potentially bias the outcome of the algorithms.^19^ Future work will investigate the mechanisms by which a model’s design, data, and deployment may lead to disparities in AD. Developing a fairness-constrained model may be one avenue to tackle the fairness challenges highlighted in this paper. This paper highlights the potential for unfairness in ML-based AD prediction modeling and highlights the importance of devoting attention to mitigating bias and advancing health equity.

## Data Availability

All data produced in the present work are contained in the manuscript

## Supplement

### Training details

All experiments in this study were conducted on One Nvidia RTX 3090 GPU, one Inter i9-12900F CPU with 16 cores and 32G RAM. Pytorch 1.0 and Python 2.7 were used to define all models and training procedures.

For logistic regression, we used the implementation of logistic regression in sklearn library^48^ for a three-class classification problem. We used L2 penalty (ridge regression) of weights, 1000 iterations, and the default solver lbfgs^49^ to learn the weights. To train a SVM model, we followed the work from Nguyen.^35^ Since SVM accepts fixed length feature vectors and it cannot handle subjects with different number of inputs timepoints. We trained different SVM models using 1 to 4 input timepoints (spaced 6 months apart) to predict the future observations. We trained 40 SVM models on four input timepoints (1, 2, 3 or 4 input timepoints) to predict clinical diagnosis as outcome for 10 future predictions (6, 12, 18,…, 60 months), in which 4×10 = 40 SVM models. The four timepoints were validated as the best settings in Nguyen’s work.^35^ The maximum iteration for training SVM is set to 10^5^. The SVM model utilized the radial basis function kernel, and the process of tuning hyperparameters remained consistent with the approach described in Nguyen’s paper ^35^. To train the RNN model, we set batch size as 128 and epoch number aS200. We use Adam^50^ as the optimizer with learning rate of 5 × 10^−4^, the value of β1 as 0.9 and β2 as 0.999, and weight decay as 5 × 10^−7^to avoid overfitting. As in Nguyen’s paper ^35^, we used an unweighted sum of cross-entropy loss for categorical variable (diagnosis stage) and MAE loss for the continuous variables. The selected model was determined by the best accuracy on validation data set. As the focus of this paper is on assessing the fairness in machine learning models on predicting AD as opposed to risk prediction model development, we do not report details of the predictors and model performance during the training phase in detail. Further description of variables and model performance can be found in TADPOLE challenge^31^ and Nguyen’s work.^35^

### Model performance

Following the same evaluation of model performance in Nguyen’s work,^35^ diagnosis classification accuracy was evaluated using the multiclass area under the operating curve (mAUC)^51^ and balanced class accuracy (BCA) metrics. The mAUC was computed as the average of three two-class AUC (AD vs not AD, MCI vs. not MCI, and CN vs not CN). For both mAUC and BCA metrics, higher values indicate better performance. The performance was evaluated by averaging the results across 10 test sets for logistic regression, SVM, and RNN. The results for the three models are shown in Table S2.

**Figure S1.**
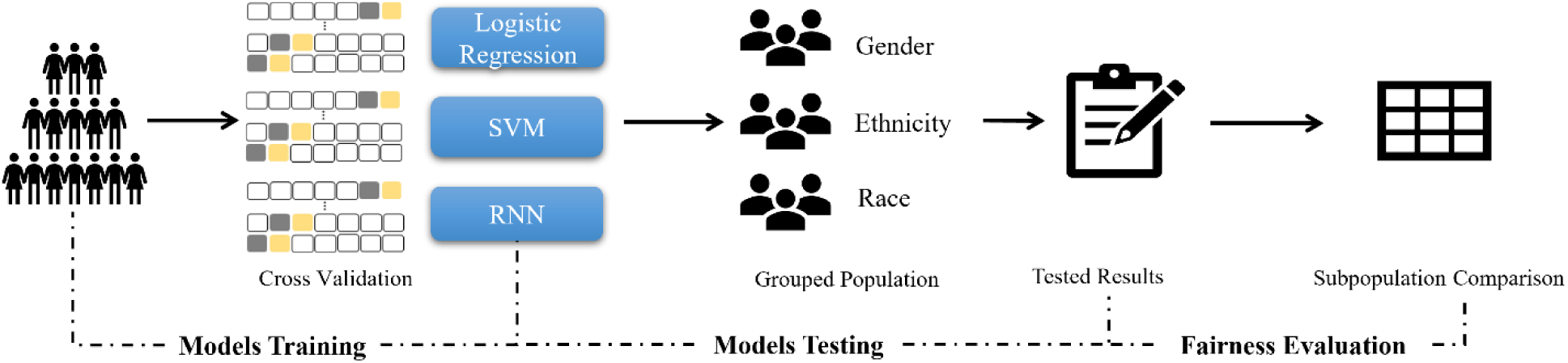
An overview of the model pipeline. Model Training: we trained three ML models using cross-validation from entire populations to predict the progression to AD; Model Testing: we tested three models across the different grouped populations, including gender, ethnicity, and race; Fairness Evaluation: we assessed the fairness metrics on test results.

**Table S1.**
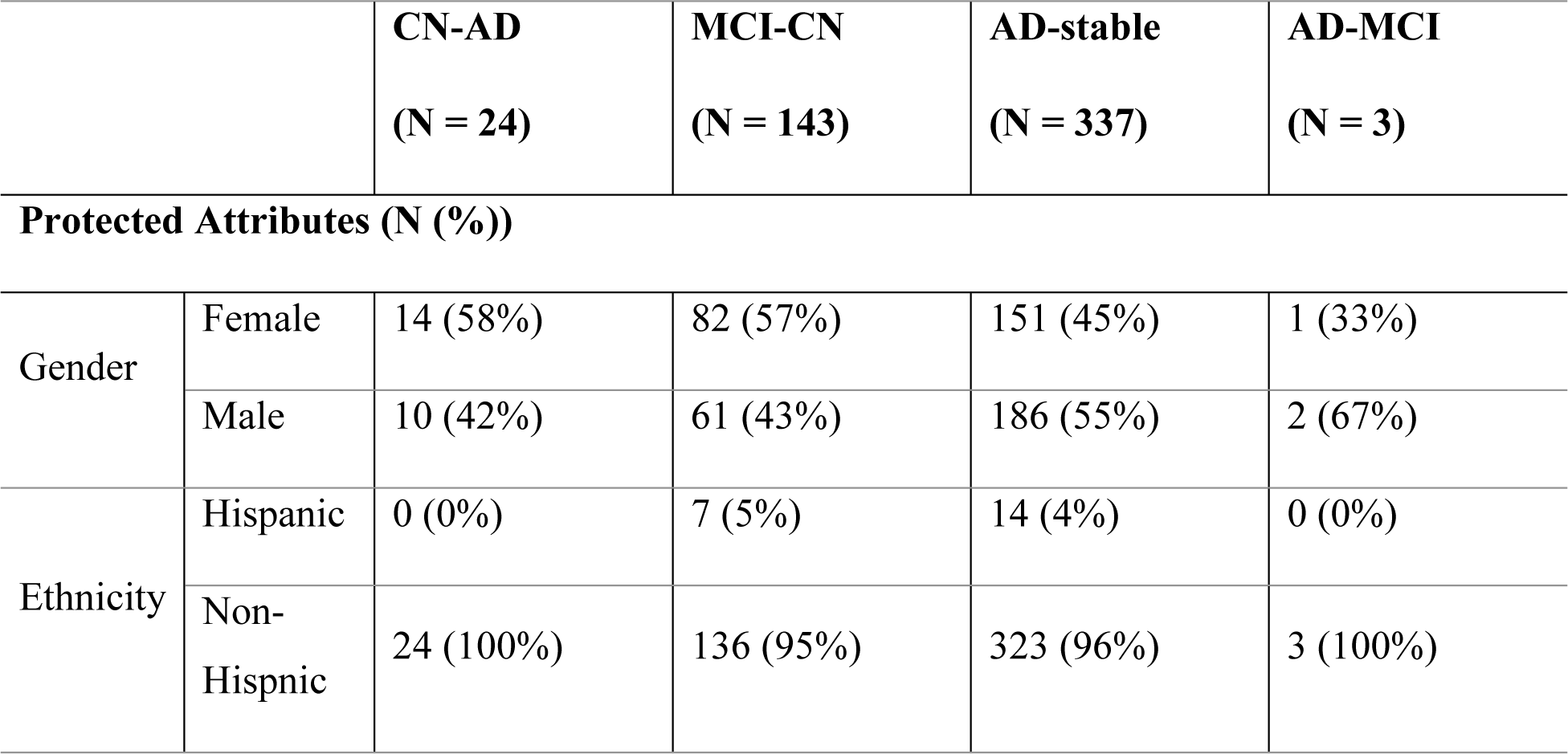
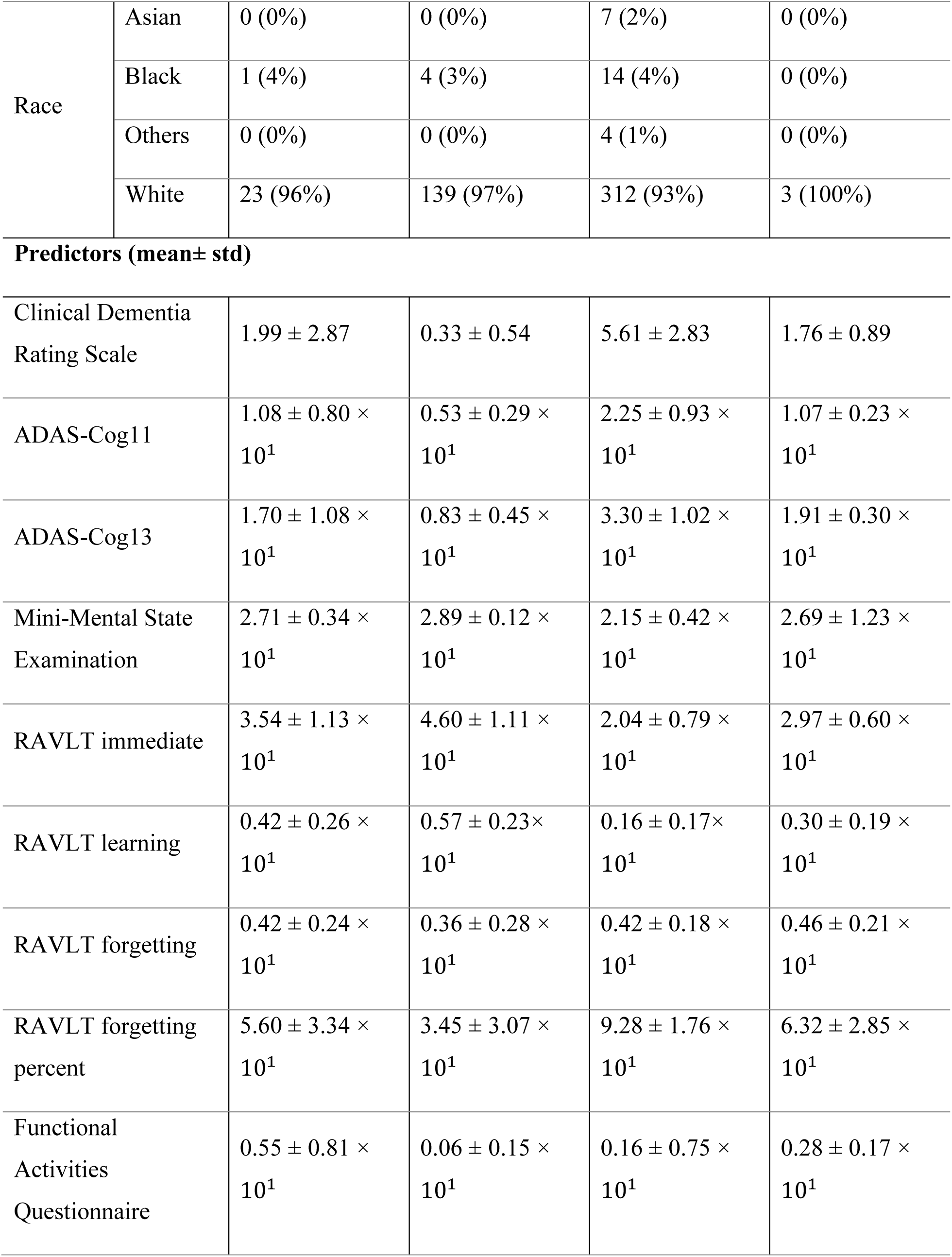
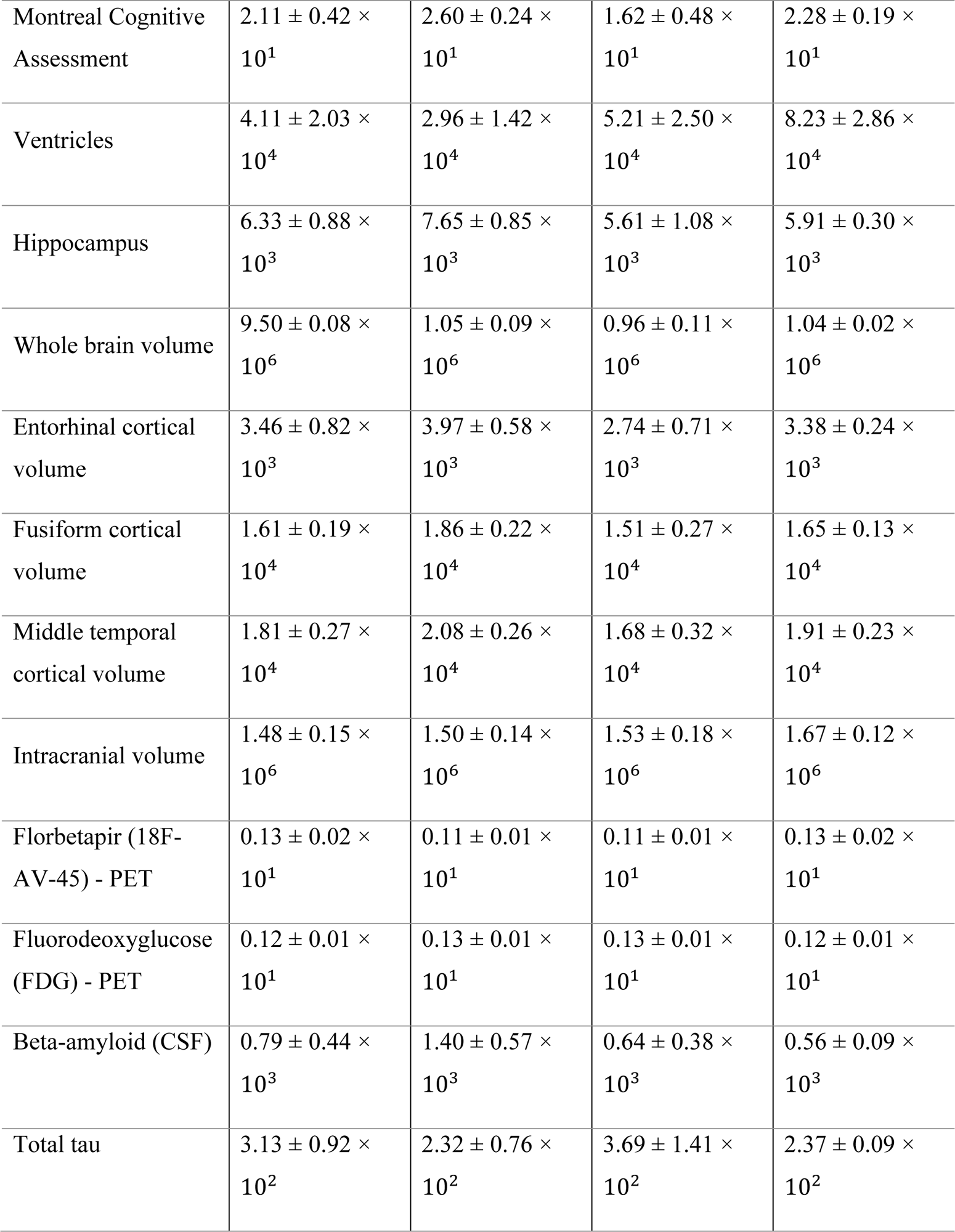
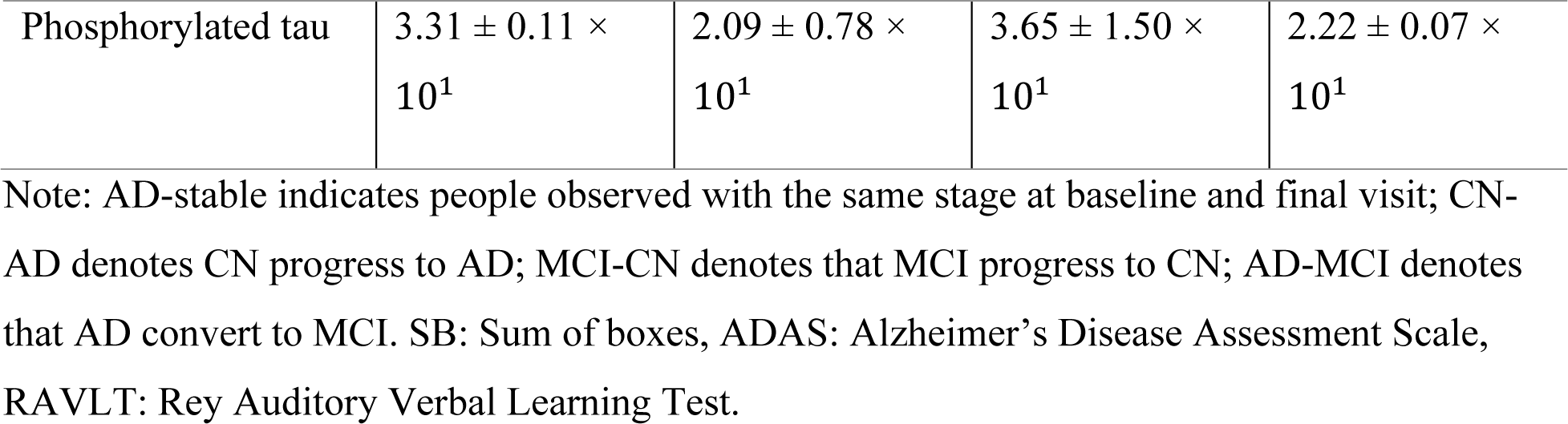
Summary statistics for protected attributes and predictor variables stratified by cognitive functioning trajectory (CN-AD, MCI-CN, AD-stable and AD-MCI) for trajectories excluded from fairness analysis due to small sample size.

|  |  | **CN-AD**<br>(N = 24) | **MCI-CN**<br>(N = 143) | **AD-stable**<br>(N = 337) | **AD-MCI**<br>(N = 3) |
| --- | --- | --- | --- | --- | --- |
| **Protected Attributes (N (%))** |  |  |  |  |  |
| Gender | Female | 14 (58%) | 82 (57%) | 151 (45%) | 1 (33%) |
|  | Male | 10 (42%) | 61 (43%) | 186 (55%) | 2 (67%) |
| Ethnicity | Hispanic | 0 (0%) | 7 (5%) | 14 (4%) | 0 (0%) |
|  | Non-Hispanic | 24 (100%) | 136 (95%) | 323 (96%) | 3 (100%) |
| Race | Asian | 0 (0%) | 0 (0%) | 7 (2%) | 0 (0%) |
|  | Black | 1 (4%) | 4 (3%) | 14 (4%) | 0 (0%) |
|  | Others | 0 (0%) | 0 (0%) | 4 (1%) | 0 (0%) |
|  | White | 23 (96%) | 139 (97%) | 312 (93%) | 3 (100%) |

**Predictors (mean± std)**
|  |  |  |  |  |
| --- | --- | --- | --- | --- |
| Clinical Dementia Rating Scale | $1.99 \pm 2.87$ | $0.33 \pm 0.54$ | $5.61 \pm 2.83$ | $1.76 \pm 0.89$ |
| ADAS-Cog11 | $1.08 \pm 0.80 \times 10^1$ | $0.53 \pm 0.29 \times 10^1$ | $2.25 \pm 0.93 \times 10^1$ | $1.07 \pm 0.23 \times 10^1$ |
| ADAS-Cog13 | $1.70 \pm 1.08 \times 10^1$ | $0.83 \pm 0.45 \times 10^1$ | $3.30 \pm 1.02 \times 10^1$ | $1.91 \pm 0.30 \times 10^1$ |
| Mini-Mental State Examination | $2.71 \pm 0.34 \times 10^1$ | $2.89 \pm 0.12 \times 10^1$ | $2.15 \pm 0.42 \times 10^1$ | $2.69 \pm 1.23 \times 10^1$ |
| RAVLT immediate | $3.54 \pm 1.13 \times 10^1$ | $4.60 \pm 1.11 \times 10^1$ | $2.04 \pm 0.79 \times 10^1$ | $2.97 \pm 0.60 \times 10^1$ |
| RAVLT learning | $0.42 \pm 0.26 \times 10^1$ | $0.57 \pm 0.23 \times 10^1$ | $0.16 \pm 0.17 \times 10^1$ | $0.30 \pm 0.19 \times 10^1$ |
| RAVLT forgetting | $0.42 \pm 0.24 \times 10^1$ | $0.36 \pm 0.28 \times 10^1$ | $0.42 \pm 0.18 \times 10^1$ | $0.46 \pm 0.21 \times 10^1$ |
| RAVLT forgetting percent | $5.60 \pm 3.34 \times 10^1$ | $3.45 \pm 3.07 \times 10^1$ | $9.28 \pm 1.76 \times 10^1$ | $6.32 \pm 2.85 \times 10^1$ |
| Functional Activities Questionnaire | $0.55 \pm 0.81 \times 10^1$ | $0.06 \pm 0.15 \times 10^1$ | $0.16 \pm 0.75 \times 10^1$ | $0.28 \pm 0.17 \times 10^1$ |
| Montreal Cognitive Assessment | $2.11 \pm 0.42 \times 10^1$ | $2.60 \pm 0.24 \times 10^1$ | $1.62 \pm 0.48 \times 10^1$ | $2.28 \pm 0.19 \times 10^1$ |
| Ventricles | $4.11 \pm 2.03 \times 10^4$ | $2.96 \pm 1.42 \times 10^4$ | $5.21 \pm 2.50 \times 10^4$ | $8.23 \pm 2.86 \times 10^4$ |
| Hippocampus | $6.33 \pm 0.88 \times 10^3$ | $7.65 \pm 0.85 \times 10^3$ | $5.61 \pm 1.08 \times 10^3$ | $5.91 \pm 0.30 \times 10^3$ |
| Whole brain volume | $9.50 \pm 0.08 \times 10^6$ | $1.05 \pm 0.09 \times 10^6$ | $0.96 \pm 0.11 \times 10^6$ | $1.04 \pm 0.02 \times 10^6$ |
| Entorhinal cortical volume | $3.46 \pm 0.82 \times 10^3$ | $3.97 \pm 0.58 \times 10^3$ | $2.74 \pm 0.71 \times 10^3$ | $3.38 \pm 0.24 \times 10^3$ |
| Fusiform cortical volume | $1.61 \pm 0.19 \times 10^4$ | $1.86 \pm 0.22 \times 10^4$ | $1.51 \pm 0.27 \times 10^4$ | $1.65 \pm 0.13 \times 10^4$ |
| Middle temporal cortical volume | $1.81 \pm 0.27 \times 10^4$ | $2.08 \pm 0.26 \times 10^4$ | $1.68 \pm 0.32 \times 10^4$ | $1.91 \pm 0.23 \times 10^4$ |
| Intracranial volume | $1.48 \pm 0.15 \times 10^6$ | $1.50 \pm 0.14 \times 10^6$ | $1.53 \pm 0.18 \times 10^6$ | $1.67 \pm 0.12 \times 10^6$ |
| Florbetapir (18F-AV-45) - PET | $0.13 \pm 0.02 \times 10^1$ | $0.11 \pm 0.01 \times 10^1$ | $0.11 \pm 0.01 \times 10^1$ | $0.13 \pm 0.02 \times 10^1$ |
| Fluorodeoxyglucose (FDG) - PET | $0.12 \pm 0.01 \times 10^1$ | $0.13 \pm 0.01 \times 10^1$ | $0.13 \pm 0.01 \times 10^1$ | $0.12 \pm 0.01 \times 10^1$ |
| Beta-amyloid (CSF) | $0.79 \pm 0.44 \times 10^3$ | $1.40 \pm 0.57 \times 10^3$ | $0.64 \pm 0.38 \times 10^3$ | $0.56 \pm 0.09 \times 10^3$ |
| Total tau | $3.13 \pm 0.92 \times 10^2$ | $2.32 \pm 0.76 \times 10^2$ | $3.69 \pm 1.41 \times 10^2$ | $2.37 \pm 0.09 \times 10^2$ |
| Phosphorylated tau | $3.31 \pm 0.11 \times 10^1$ | $2.09 \pm 0.78 \times 10^1$ | $3.65 \pm 1.50 \times 10^1$ | $2.22 \pm 0.07 \times 10^1$ |

Note: AD-stable indicates people observed with the same stage at baseline and final visit; CN-AD denotes CN progress to AD; MCI-CN denotes that MCI progress to CN; AD-MCI denotes that AD convert to MCI. SB: Sum of boxes, ADAS: Alzheimer's Disease Assessment Scale, RAVLT: Rey Auditory Verbal Learning Test.

**Table S2.** Prediction performance averaged across 10 test sets.

| | mAUC<br>(mean ± SD) | BCA<br>(mean ± SD) |
| --- | --- | --- |
| LR | $0.916 \pm 0.017$ | $0.825 \pm 0.023$ |
| SVM | $0.921 \pm 0.011$ | $0.831 \pm 0.021$ |
| RNN | $0.949 \pm 0.008$ | $0.891 \pm 0.017$ |

In each cell, the two numbers represent the mean and standard deviation derived from 10 tests. mAUC = multiclass area under the operating curve; BCA = balanced class accuracy. LR= logistic regression; SVM = Support Vector Machine; RNN = recurrent neural networks.

**Figure S2.**
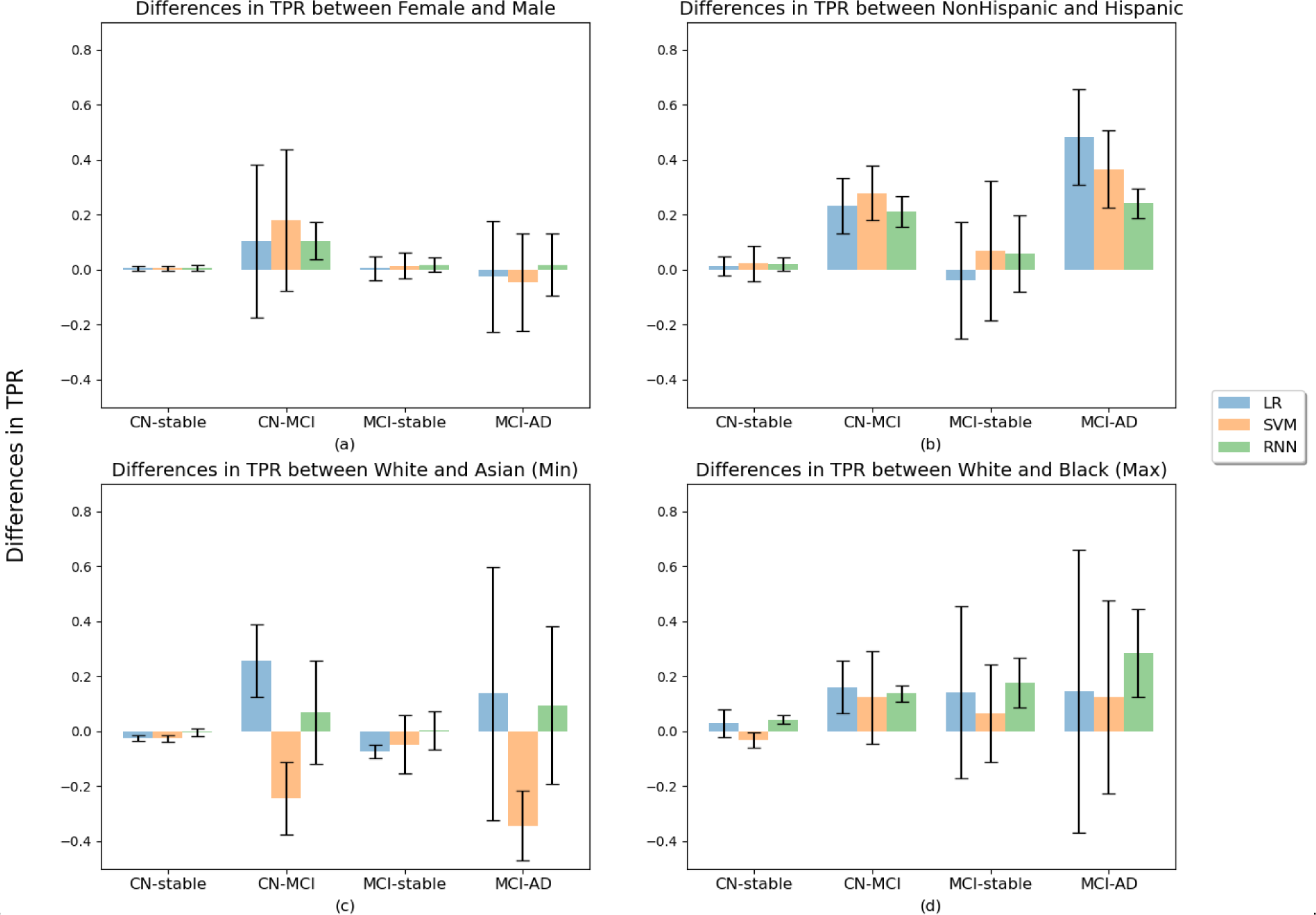
Absolute differences in TPR across groups defined by the three protected attributes. For gender, the difference in TPR is between male and female. For ethnicity, the difference in TPR is reported between Non-Hispanic and Hispanic. For race, since there are four groups, we first compute all pairwise differences with the “White” group which we considered as a reference as it had the largest sample size. We then report the minimum differences in TPR, which resulted from the contrast between the White and Asian groups, and the maximum differences, which resulted from comparing the White and Black groups. Bars represent mean values across 10 test sets and error bars represent a corresponding standard deviation of the 10 mean values.

**Figure S3.**
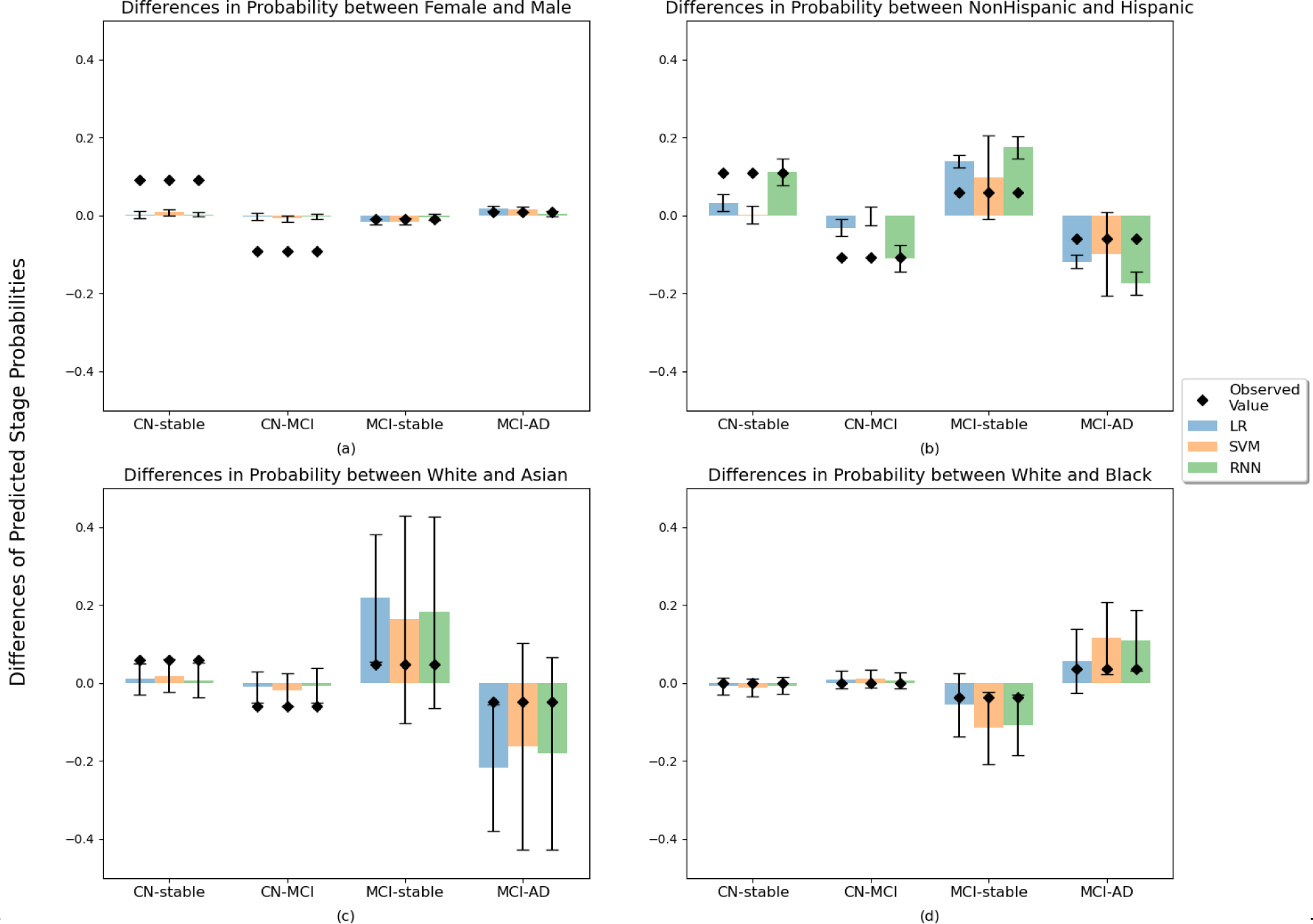
Differences of predicted progression probabilities between groups of each protected attribute with three evaluated models. The results are averaged over 10 test sets using predictions from the LR, SVM, and RNN models. Bars represent the mean values across 10 test sets and error bars represent a corresponding standard deviation of the 10 mean values. Dots represent the average values of differences of the empirical probability of each trajectory stratified by demographic subgroup on 10 test sets.

## Notes

### Competing Interest Statement

The authors have declared no competing interest.

### Funding Statement

Research reported in this publication was supported by the National Institutes of Health under award number R21AG075574.

